# 5-Alpha-Reductase Inhibitors Reduce Remission Time of COVID-19: Results From a Randomized Double Blind Placebo Controlled Interventional Trial in 130 SARS-CoV-2 Positive Men

**DOI:** 10.1101/2020.11.16.20232512

**Authors:** Flávio Adsuara Cadegiani, John McCoy, Carlos Gustavo Wambier, Andy Goren

**Affiliations:** Department of Clinical Endocrinology, Federal University of São Paulo Medical School, Sao Paulo, Brazil; Applied Biology, Inc. Irvine, CA, USA; Department of Dermatology, Alpert Medical School of Brown University, Providence, RI, USA

**Keywords:** COVID-19, SARS-CoV-2, androgen receptor, androgenetic alopecia, anti-androgen therapy, transmembrane protease serine 2, TMPRSS2, Dutasteride, Finasteride, 5-alpha reductase

## Abstract

**Background:** SARS-CoV-2 entry into type II pneumocytes is depended on the TMPRSS2 proteolytic enzyme. The only known promoter of TMPRSS2 in humans is an androgen response element. As such, androgen sensitivity may be a risk factor for COVID-19. Previously, we have reported a retrospective cohort analysis demonstrating the protective effect of 5-alpha-reductase inhibitors (5ARis) in COVID-19. Men using 5ARis were less likely to be admitted to the ICU than men not taking 5ARis. Additionally, men using 5ARis had drastically reduced frequency of symptoms compared to men not using 5ARis in an outpatient setting. Here we aim to determine if 5ARis will be a beneficial treatment if given after SARS-CoV-2 infection.

**Methods:** A double-blinded, randomized, prospective, investigational study of dutasteride for the treatment of COVID-19 (NCT04446429). Subjects confirmed positive for SARS-CoV-2 were treated in an outpatient setting. Endpoints for the study were remission times for a predetermined set of symptoms: fever or feeling feverish, cough, shortness of breath, sore throat, body pain or muscle pain/ache, fatigue, headache, nasal congestion, nasal discharge (runny nose), nausea or vomiting, diarrhea, loss of appetite, and loss of taste or smell (Ageusia or Anosmia).

**Results:** A total of 130 SARS-CoV-2 positive men were included in the study, 64 subjects in the dutasteride arm and 66 subjects in the placebo-controlled group. The differences in the average remission times for fatigue and anosmia or ageusia was statistically different between the groups (5.8 versus 10.1 days for fatigue and 7.3 versus 13.4 days for anosmia or ageusia, in dutasteride and placebo groups, respectively), however, the total remission time was significantly reduced for the men given dutasteride; 9.0 days versus 15.6 days in the placebo group (p < 0.001). Excluding loss of taste and smell the average total remission time was 7.3 days in the dutasteride group versus 11.7 in the placebo arm (p < 0.001).

**Conclusion:** The total remission time for men using 5ARis was significantly reduced compared to men not taking 5ARis.

## Introduction

Early in the COVID-19 pandemic, reports from Wuhan, China demonstrated that the infectivity and severity of the disease disproportionately affects men. Of patients sampled in the early stages of the outbreak 42% were female versus 58% male.^1^ Now that the disease has progressed to the majority of countries across the globe, the trend has been demonstrated many times over; men are more likely to be infected, more likely to have severe disease, and have a greater case fatality rate compared to women.^2^ Lifestyle differences and gender-biased comorbidities, e.g., incidence of smoking and hypertension, have been suggested as contributing to this gender discrepancy,^2^ however, definitive proof of these associations is lacking. We have previously published several communications suggesting that the male bias in COVID-19 disease severity may be linked to androgens.^3,4^

SARS-CoV-2 entry into type II pneumocytes is dependent on modification of a viral spike protein by the transmembrane protease, serine 2 (TMPRSS2) expressed on the surface of human cells.^5^ The only known promoter of the TMPRSS2 gene in humans is an androgen response element located in the 5’ promoter region.^6^ It would follow that reducing the expression of TMPRSS2 by blocking the androgen receptor would decrease SARS-CoV-2 entry into human cells. Recently, we have published several observation studies linking the androgen-mediated phenotype of androgenetic alopecia (AGA) to COVID-19 disease severity.^3,4^ In a cohort of 122 men hospitalized with COVID-19, 79% were diagnosed with AGA compared to the expected prevalence of 31-53% in aged matched controls of similar ethnicity.^3^ Additionally, a recent publication has demonstrated that COVID-19 disease severity was directly correlated with AGA progression; men with higher Hamilton-Norwood stages were more likely to experience severe disease and death.^7^

Further evidence connecting COVID-19 to androgens has been reported in prostate cancer patients undergoing androgen depravation therapy (ADT). Montopoli et al. studied a large population of COVID-19 patients in northern Italy, observing that COVID-19 infection rates were lower in prostate cancer patients receiving ADT compared to prostate cancer patients not receiving ADT (OR 4.05; 95% CI 1.55-10.59).^8^ Other groups have suggested that polycystic ovary syndrome may also indicated increase risk of severe COVID-19 disease in women,^9^ however, clinical data supporting this hypothesis have not been produced. Finally, we have communicated that variations in the androgen receptor gene may contribute to the racial variations in case fatality rates observed in the United States.^10^ Taken together, there is growing body of evidence to support that SARS-CoV-2 infectivity is mediated by the androgen receptor and may respond to drugs that reduce androgen receptor function.

5-alpha-reductase inhibitors (5ARis) are commonly prescribed for androgenetic alopecia and benign prostatic hyperplasia; they block the conversion of testosterone to the more potent androgen, dihyrotestosterone (DHT).^11^ 5ARis are inexpensive and have, compared to other ADTs, relatively low incidence of adverse side effects. As such, they would make ideal candidates for a SARS-CoV-2 treatment. Recently, we have reported the results from two retrospective cohort analysis demonstrating the protective effect of 5-alpha-reductase inhibitors (5ARi) for men with COVID-19.^12^ In a study of 77 men hospitalized with COVID-19 we found among men taking 5ARis, 8% were admitted to the ICU compared to 58% of men not taking 5ARis (P = 0.0015). In the cohort, 5ARis were associated with reduced risk for ICU admissions RR 0.14 (95% CI: 0.02–0.94).^12^ Similarly, we have demonstrated that the frequency of COVID-19 symptoms was drastically reduced for men using 5ARis in an outpatient setting. A statistically significant (p<0.05) reduction in the frequency of 20 of the 29 clinical symptoms was observed in AGA men using 5ARis compared to AGA men not using 5ARis. For example, 38% and 2% of men presented with low-grade fever, 60% and 6% with dry cough, and 88% and 15% reported anosmia in the non-5ARi and 5ARi groups, respectively.^13^ In this communication we sought to determine if 5ARis are a beneficial treatment for COVID-19 if given after SARS-CoV-2 infection.

## Methods

### Study Design and Oversight

Potential subjects were recruited to a double-blinded, randomized, prospective, investigational study of anti-androgen treatment of COVID-19 (NCT04446429). Prospective subjects for the study presented with mild symptoms of suspected COVID-19 infection at one of five outpatient clinics (Corpometria Institute Brasilia, Brazil) and did not require admission to a hospital at the time of their visit. The study is registered (clinicaltrial.gov) and conducted with the consent and approval of the Brazilian National Ethics Committee (CAAE: 34110420.2.0000.0008). All patients admitted to the study gave informed consent.

Baseline characteristics, comorbidities, test results, and medications used were extracted from patient records. For each subject, the age, BMI (kg/m^2^), frequency and duration of medication used and the following pre-existing conditions were extracted from records: type 2 diabetes, hypertension, obesity (BMI), hypothyroidism, hypogonadism, androgenetic alopecia, asthma, and chronic obstructive pulmonary disease (COPD). Data were extracted by the principal investigator and managed by the study director.

### Study Population and Covariates

Males being screened for inclusion in a double-blinded, randomized, prospective, interventional study of anti-androgen treatment of COVID-19 were recruited through social media as well as a mailing list of 10,900 men from the Brazilian health care system registry. Screening of potential subjects was conducted at five outpatient clinics (Corpometria Institute Brasilia, Brazil) at which nasopharyngeal swabs were collected by trained medical personal. SARS-CoV-2 status was laboratory confirmed by real-time reverse transcription polymerase reaction testing (Automatized Platform, Roche, USA) following the Cobas SARS-CoV-2 rtPCR kit test protocol.

### Study Outcomes

Endpoints for the study were remission times for the following symptoms: fever or feeling feverish, cough, shortness of breath, sore throat, body pain or muscle pain/ache, fatigue, headache, nasal congestion, nasal discharge (runny nose), nausea or vomiting, diarrhea, loss of appetite, and loss of taste or smell (Ageusia or Anosmia). The average remission time (days) of each symptom was tabulated for each group. Additionally, the total remission time was calculated for each group. Finally, patients were asked to rate their disease severity at various times before and after treatment began. The times used were Day -7 to -4, Day -3 to -1, Day 0 (start of treatment), Day 1, Day 2, Day 3, Day 7, Day 14, Day 21, Day 30, and Day 60. Subjects were asked to rate their health on a scale from 0 to 100, 100 being completely heathy and 0 being their worst day of COVID-19.

### Statistical Analysis

Medical history, concomitant medications and lifestyle characteristics of COVID-19 patients were tabulated based for each group: age, BMI, hypertension, myocardial infarction, stroke, heart failure, lipid disorders, diabeties, pre-diabeties, obesity, asthma, COPD, cancer, benign prostatic hyperplasia, prostate cancer, chronic renal disease, liver fibrosis/cirrhosis, clinical depression, anxiety, ADHD, insomnia, hypogonadism, hypothyroidism, and autoimmune disorders as well as indicated medications. For each clinical symptom, the average remission time was calculated for each group. The Fisher exact test was used to compare the remission times for clinical symptoms between COVID-19 patients using 5ARis and COVID-19 patients not using 5ARis. Statistical significance was set at p<0.05. XLSTAT version 2020.3.1.1008 (Addinsoft, Inc.) was used to perform all statistical analysis.

## Results

There were 130 SARS-CoV-2 positive men included in the trial. 64 men were assigned to the investigational arm and 66 men were assigned to the placebo group. Average interval between first symptoms and beginning of treatment was 4.26 and 4.25 days for investigational and placebo groups, respectively (p = n/s). Baseline characteristics of the two study groups are displayed in **Table 1**.

**Table 1.** Characteristics of the study populations. Shown are the characteristic of the two arms of the study Congestive heart failure = CHF; type 2 diabetes *mellitus* = T2DM, chronic obstructive pulmonary disorder = COPD, chronic kidney disease = CKD, attention deficit hyperactivity disorder = ADHD, benign prostate hyperplasia = BPH, angiotensin converting enzyme inhibitors = ACEi; angiotensin-2 receptor blockers = ARB; calcium channel blocker = CCB; glucagon-like peptide-1 receptor analogues = GLP1Ra; sodium-glucose cotransporter-2 inhibitors = SGLT2i; dipeptidyl-peptidase 4 inhibitors = DPP4i; progesterone = P; estradiol = E; gonadotropin release hormone = GnRH; selective estrogen receptor modulators = SERM; non-steroidal antiandrogen = NSAA; selective serotonin reuptake inhibitors = SSRI; central nervous system = CNS; Bacillus Calmette-Guérin = BCG;

|  | Placebo |  | Dutasteride |  |
| --- | --- | --- | --- | --- |
|  | AVG | STD | AVG | STD |
| AGE (y/o) | 42.0 | 11.5 | 42.0 | 12.6 |
| WEIGHT (kg) | 85.5 | 11.6 | 85.5 | 9.8 |
| HEIGHT (m) | 1.8 | 0.1 | 1.8 | 0.1 |
| BMI (Kg/m2) | 26.5 | 2.8 | 26.5 | 3.2 |
| N | 66.0 |  | 64.0 |  |
|  | N | % | N | % |
| Obesity | 8 | 12 | 9 | 14 |
| Married? | 47 | 71 | 43 | 67 |
| Live alone? | 12 | 18 | 13 | 20 |
| Hypertension | 18 | 27 | 15 | 23 |
| Mi | 1 | 2 | 0 | 0 |
| Stroke | 0 | 0 | 0 | 0 |
| Hf | 0 | 0 | 1 | 2 |
| Lipid disorders | 14 | 21 | 16 | 25 |
| Other cardiac dysfunctions | 0 | 0 | 3 | 5 |
| Diabetes | 7 | 11 | 7 | 11 |
| Pré-dm2 | 11 | 17 | 9 | 14 |
| Asthma | 1 | 2 | 0 | 0 |
| COPD | 0 | 0 | 0 | 0 |
| Chronic renal dis. | 0 | 0 | 0 | 0 |
| Liver fibrosis/chirrosis | 1 | 2 | 0 | 0 |
| Clinical depression | 5 | 8 | 4 | 6 |
| Anxiety | 13 | 20 | 12 | 19 |
| ADHD | 5 | 8 | 6 | 9 |
| Insomnia | 4 | 6 | 6 | 9 |
| Hypothyroidism | 3 | 5 | 4 | 6 |
| Autoimmune disorders | 1 | 2 | 0 | 0 |
| Cancer | 0 | 0 | 0 | 0 |
| Hypogonadism | 13 | 20 | 11 | 17 |
| BPH | 2 | 3 | 3 | 5 |
| **DRUGS** |  |  |  |  |
| Beta-blocker | 3 | 5 | 3 | 5 |
| ECAi | 4 | 6 | 1 | 2 |
| ARB | 14 | 21 | 14 | 22 |
| Loop diur. | 3 | 5 | 0 | 0 |
| Thiazide diuretics | 4 | 6 | 5 | 8 |
| CCB | 2 | 3 | 4 | 6 |
| Statins | 14 | 21 | 15 | 23 |
| **Others** | 0 | 0 | 1 | 2 |
| **Aspirin** | 0 | 0 | 2 | 3 |
| **Clopidogrel** | 0 | 0 | 0 | 0 |
| **Warfarin** | 0 | 0 | 0 | 0 |
| **Xa factor inhibitors** | 0 | 0 | 1 | 2 |
| **Direct thrombin inhibitors** | 0 | 0 | 0 | 0 |
| **Heparins** | 0 | 0 | 0 | 0 |
| **Metformin** | 16 | 24 | 14 | 22 |
| **GLP1R analogue** | 4 | 6 | 8 | 13 |
| **SGLT2 inhibitors** | 8 | 12 | 7 | 11 |
| **DPP4 inhibitors** | 3 | 5 | 2 | 3 |
| **Sulfonylureas** | 1 | 2 | 0 | 0 |
| **Glitazone** | 0 | 0 | 0 | 0 |
| **Acarbose** | 0 | 0 | 0 | 0 |
| **Insulin** | 0 | 0 | 0 | 0 |
| **Orlistat** | 4 | 6 | 5 | 8 |
| **Levothyroxine** | 3 | 5 | 4 | 6 |
| **Liothyronine** | 1 | 2 | 0 | 0 |
| **Testosterone** | 11 | 17 | 9 | 14 |
| **Aromatase inhibitors or SERMs** | 2 | 3 | 2 | 3 |
| **Hipnotics** | 4 | 6 | 5 | 8 |
| **Selective serotonin reuptaker inhibitors (SSRI)** | 10 | 15 | 9 | 14 |
| **Other antidepressants and humor stabilizers** | 7 | 11 | 6 | 9 |
| **Benzodiazepines** | 2 | 3 | 2 | 3 |
| **Atypical antipsychotics** | 4 | 6 | 3 | 5 |
| **CNS stimulants** | 5 | 8 | 8 | 13 |
| **Alpha-1 adren. Blockers** | 2 | 3 | 3 | 5 |
| **Gnrh analogues and inh., NSAA, others** | 0 | 0 | 0 | 0 |
| **Erectile dysfunction** | 2 | 3 | 1 | 2 |
| **Omega-3** | 3 | 5 | 1 | 2 |
| **Vitamin D** | 17 | 26 | 17 | 27 |
| **Zinc** | 6 | 9 | 7 | 11 |
| **Biotin** | 1 | 2 | 0 | 0 |
| **Vitamin C** | 8 | 12 | 8 | 13 |
| **Multivitamin** | 2 | 3 | 2 | 3 |
| **BCG** | 64 | 97 | 63 | 98 |
| **INFLUENZA (in 2020)** | 20 | 30 | 21 | 33 |
| **PNEUMOCOCCAL (since 2017)** | 4 | 6 | 8 | 13 |
| **CURRENT SMOKING** | 4 | 6 | 6 | 9 |
| **REGULAR PHYSICAL ACTIVITY** | 41 | 62 | 42 | 66 |
| **ADDITIONAL COVID TREATMENTS** |  |  |  |  |
| Ivermectin | 7 | 11 | 6 | 9 |
| Hydroxychlorouquine | 4 | 6 | 3 | 5 |
| Xa FACTOR INHIBITORS | 9 | 14 | 6 | 9 |
| Enoxaparin | 4 | 6 | 4 | 6 |
| Glucocorticoids | 7 | 11 | 8 | 13 |
| Vitamin c | 4 | 6 | 5 | 8 |
| Zinc | 3 | 5 | 4 | 6 |
| Vitamin d | 1 | 2 | 3 | 5 |
| Colchicine | 0 | 0 | 0 | 0 |

**None of the parameters was statistically significant between groups.**

**Table 2** reports the average remission times for clinical symptoms in men taking 5ARis versus men not taking 5ARis. In most of the symptoms recorded, the average remission time was not significantly reduced. The average remission time for fatigue was 5.8 days in the dutasteride group versus 10.1 days in the placebo group (p < 0.001). The average remission time for loss of taste or smell was 7.3 days in the dutasteride group versus 13.4 days in the placebo group (p < 0.001). Total remission for all symptoms was statistically significantly reduced. Total remission time was 9.0 days versus 15.6 days in the placebo group (p < 0.001). After excluding loss of taste and smell, the average total remission time was 7.3 days in the dutasteride group versus 11.7 in the placebo arm (p < 0.001).

**Table 2.** Clinical symptom remission of the study populations. Shown are the average remission times for each clinical symptoms of COVID-19 men using 5-alpha-reductase inhibitors (5ARi) compared to placebo.

|  | **<u>Duration of Symptoms (Days)</u>** |  |  |
| --- | --- | --- | --- |
|  | Placebo<br>(n=66) | Dutasteride<br>(n=64) | p |
| **Cough** | 3.2 | 3.1 | 0.839 |
| **Shortness of Breath** | 0.9 | 0.3 | 0.111 |
| **Fatigue** | 10.1 | 5.8 | < 0.001 |
| **Loss of Taste or Smell (Ageusia or Anosmia)** | 13.4 | 7.3 | < 0.001 |
| **Remission Time Minus Loss of Taste or Smell** | 11.7 | 7.3 | < 0.001 |
| **Total Remission Time** | 15.6 | 9.0 | < 0.001 |

**Figure 1** displays the average results from the patient reported outcome rating disease severity at various times before and after treatment began. The level of recovery was significantly improved in the dutasteride group compared to placebo group in Days 1, 2, 3 and 7 (p < 0.001 for all days).

**Figure 1.**
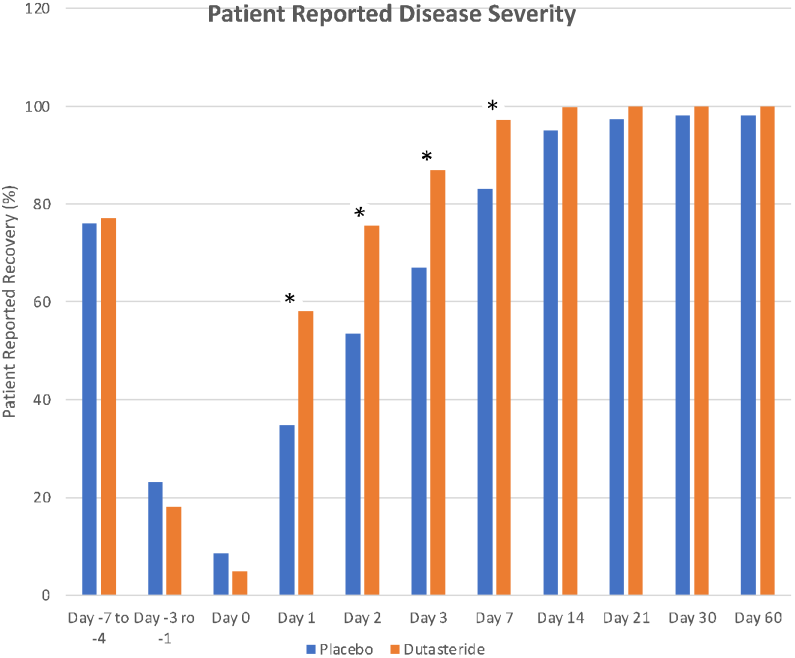
Patient reported disease severity at various times before and after treatment. Day 0 denotes the start of treatment. Patients were asked to rate their health on a scale from 0 to 100, 100 being completely heathy and 0 being their worst day of COVID-19.

## Discussion

Men infected with SARS-CoV-2 have an increased risk of severe COVID-19 disease compared to women.^2^ A multitude of factors may contribute to the gender disparity,^2^ however, evidence is mounting^3,4,7,8^ to support that androgens, the defining male hormones, may be involved in the regulation of COVID-19 severity. Concurrently, the mechanism of action is likely androgen receptor regulation of the expression of the TMPRSS2 enzyme, one of the enzymes utilized by SARS-CoV-2 to enter type II pneumocytes in human lungs.^5^

Androgens are both circulating and produced in tissue. Elevated tissue DHT is implicated in androgenetic alopecia (AGA), benign prostatic hyperplasia, and prostate cancer. In a previous communication, we reported that in a cohort of 122 hospitalized COVID-19 male patients, 79% suffered from androgenetic alopecia.^3^ Similarly, Montopoli et al.,^8^ observed that men utilizing ADT for prostate cancer were less likely to suffer severe COVID-19 disease. These observations led us to study the effect of anti-androgen therapy on COVID-19 outcomes.

Here we demonstrate in a randomized, double-blinded, placebo controlled interventional study that men treated with 5ARis, commonly used to treat AGA and benign prosthetic hyperplasia, display statistically significant improvements in COVID-19 disease remission in an outpatient setting. We are currently conducting a randomized, double-blinded, placebo controlled interventional study with a novel anti-androgen (proxalutamide) in the treatment of COVID-19 (NCT04446429).

## Data Availability

The supplement data is available under request to FAC.

## Abbreviations

5ARi: 5-alpha reductase inhibitors
TMPRSS2: Transmembrane protease, serine 2
AGA: Androgenetic alopecia

